# Fraction of COVID-19 hospitalizations and deaths attributable to chronic diseases

**DOI:** 10.1101/2021.04.12.21255346

**Authors:** Francisco Reyes-Sánchez, Ana Basto-Abreu, Rossana Torres-Alvarez, Francisco Canto-Osorio, Romina González-Morales, Dwight Dyer, Ruy López Ridaura, Christian Arturo Zaragoza Jiménez, Juan A. Rivera, Tonatiuh Barrientos-Gutiérrez

## Abstract

**Aim:** To estimate the fraction of hospitalizations and deaths from COVID-19 attributable to chronic diseases due to poor nutrition and smoking in Mexico.

**Methods:** We used data from the Mexican surveillance system of COVID-19. We considered six chronic diseases (obesity, COPD, hypertension, diabetes, cardiovascular disease, and chronic kidney disease) to define a multimorbidity variable: no diseases, 1 disease, 2 diseases, or 3 or more diseases. We calibrated the database using bias quantification methods to consider the undiagnosed cases of chronic diseases. To estimate the risks of hospitalization and death due to chronic diseases, we fitted Poisson regression models with robust standard errors, adjusting for possible confounders. Using these risks, we calculated attributable fractions using the population attributable fraction (PAF).

**Results:** Chronic diseases accounted for to 25.4% (24.8%, 26.1%), 28.3% (27.8%, 28.7%) and 15.3% (14.9%,15.7%) of the hospitalizations among adults below 40 years, 40 to 59, and 60 years and older respectively (95% CI). For COVID-19-related deaths, 50.1% (48.6%, 51.5%), 40.5% (39.7%, 41.3%), and 18.7% (18.0%, 19.5%) were attributable to chronic diseases in adults under 40 years, 40 to 59, and 60 years and older, respectively.

**Conclusion:** Chronic diseases linked to malnutrition and tobacco use contributed to a higher burden of hospitalization and deaths from COVID-19 in Mexico, particularly among younger adults. Medical and structural interventions to curb chronic disease incidence and facilitate disease control are urgently needed.

## Introduction

Chronic diseases increase the risk of severity and mortality associated to COVID-19. Meta-analytical evidence shows people with chronic obstructive pulmonary disease, diabetes mellitus, hypertension and cardiovascular disease present higher risk of developing severe COVID-19 [1]. Similar findings have been reported for mortality, where obesity, diabetes mellitus, hypertension, and cardiovascular disease have been consistently associated with an increased risk of death [2].

Etiological studies have studied the association between COVID-19 and chronic diseases, producing risk estimates. Yet, the proportion of COVID-19 cases and complications attributable to chronic diseases remains understudied. In particular, the fraction of COVID-19 deaths attributable to chronic diseases could shed light into the large heterogeneity in fatality rates observed across countries [3]. By integrating the relative risk and the frequency of the exposure in the population, the population attributable fraction (PAF) provides an estimate of the burden of disease attributable to the exposure [4]. In Mexico, prior PAF estimates have shown that 1.1%, 14.3% and 16.8% of deaths from COVID-19 were attributable to diabetes, hypertension, and obesity, respectively among ambulatory patients [5]. To estimate PAFs for different risk factors is very informative, yet, it is insufficient to capture the total burden of multimorbidity [6].

Few studies to date have estimated the impact of multiple chronic diseases or multimorbidity on COVID-19 [7–9]. Estimating the PAF for multimorbidity is challenging, as the PAFs for each disease should not be simply added, given their potential overlap [6]. A second challenge is misclassification bias, considering that a large proportion of the population who suffer silent chronic conditions may not have been diagnosed. For example, in Mexico 40% of the people with hypertension are unaware of their condition [10]. Most studies rely on self-reported disease diagnosis, misclassifying a large proportion of the population with chronic conditions. To our knowledge, no study has attempted to estimate the impact of this bias on COVID-19 attributable fractions.

We aimed to estimate the fraction of hospitalizations and deaths from COVID-19 that are attributable to multimorbidity associated with poor nutrition and smoking in Mexico. We used a multimorbidity approach, based on the number of chronic diseases experienced by a single individual, considering obesity, chronic obstructive pulmonary disease (COPD), hypertension, diabetes, cardiovascular disease, and chronic kidney disease. We also implemented bias quantification methods to consider the impact of miss-classification induced by self-reported data.

## Methods

Data was obtained from the publicly available COVID-19 registry provided by the Health Ministry of Mexico [11]. The surveillance system contains information on all individuals who fulfilled the definition of suspected case by the Health Secretary: at least one major symptom (cough, fever, dyspnea or headache) and a minor symptom (myalgia, arthralgia, odynophagia, chills, chest pain, rhinorrhea, anosmia, dysgeusia, conjunctivitis) [12]. The registry used is based on the national hospital and sentinel surveillance systems and more details can be found elsewhere [13]. Briefly, patients with a diagnosis of a severe acute respiratory infection are hospitalized and all submitted to testing using reverse transcription polymerase chain reaction (PCR). Patients with a diagnosis of an acute respiratory infection are treated as outpatient and 10% of them are selected to provide a sample for PCR analysis. The 10% subsample was established in February 2020 but has increased over time. Data available includes all clinical and epidemiological information obtained at the time of registry. PCR results and deaths are updated with varying time delays, but other clinical evolution variables are not updated, such as the hospitalization of an initially ambulatory patient, discharging day, intubation, development of pneumonia, or treatment at the intensive care unit.

### Data processing

For this analysis, we included all individuals in the surveillance system registered during the community transmission phase of the pandemic (started March 23^rd^, with last update December 7^th^, 2020), with a sample size of 2,984,902 subjects. Subjects with missing values in obesity, hypertension, diabetes, chronic obstructive pulmonary disease (COPD), cardiovascular disease, and chronic kidney disease were excluded (n = 12,691). Then, we excluded people with negative (n = 1,368,252), pending or inadequate test results (n = 487,037), subjects under 20 years of age (n= 47,831), subjects that sought medical attention between November 24 to December 7 (to allow sufficient time to update PCR results or deaths in the database) (n= 48,852), and subjects that reported implausible delay in seeking medical attention (n= 13,698): reported 0 days from symptom onset to healthcare admission but were admitted with a serious condition (death, pneumonia or intensive care unit), because this would be implausible. The final sample presented 1,006,541 individuals who were positive to RT-PCR for COVID-19.

### Main Outcomes

Two outcomes were defined for this analysis: 1) Hospitalization, refers to a COVID-19 case that required inpatient care; 2) Death, defined as the death of a person with COVID-19 as recorded in the database.

### Dependent variable

Six diseases associated with poor nutrition or smoking were considered: obesity, COPD, hypertension, diabetes, cardiovascular disease, and chronic kidney disease. These diseases were selected given their prevalence and the wide range of interventions available to reduce their impact through public policy or individual-level interventions [14–17]. Information was self-reported as obtained by the medical unit’s epidemiologist when the person first sought medical attention. Then, we constructed our main dependent variable, defined as *multimorbidity* with four categories: individuals with no diseases, one disease, two diseases, or three or more diseases. Since chronic diseases information was self-reported, we conducted a quantitative bias analysis to adjust for undiagnosed cases.

### Quantitative bias analysis

To conduct the quantitative bias analysis, we estimated the proportion of undiagnosed cases by age group for diabetes and hypertension. For that, we used ENSANUT 2016 self-reported data against a gold standard (HbA1c and fasting glucose for diabetes, and systolic and diastolic blood pressure for hypertension) as a validation source [18]. The question used to assess self-reported prior medical diagnosis of diabetes or hypertension in ENSANUT-2016 was similar to the question used in the COVID-19 case-assessment format: “Has a doctor ever diagnosed you with [name of chronic disease]?”. Sensibility and specificity from ENSANUT 2016 are summarized in Table 1. Specificity was 100% because we assumed that all individuals answering “yes” in the self-reported question correctly reported their condition; due to effect of treatment (HbA1c or BP could be under control), no means exist to validate their answer.

**Table 1.** Sensitivity and specificity parameters used in the estimation of adjusted totals.

| **Age group** | **Diabetes** |  | **Hypertension** |  |
| --- | --- | --- | --- | --- |
|  | *Sensitivity (%)* | *Specificity(%)* | *Sensitivity(%)* | *Specificity(%)* |
| *20-39 years* | 61.4 | 100.0 | 48.7 | 100.0 |
| *40-60 years* | 66.5 | 100.0 | 61.8 | 100.0 |
| *60 years or older* | 76.5 | 100.0 | 64.4 | 100.0 |

To estimate the adjusted prevalence of diabetes or hypertension by age groups, we used the following formula [19]:

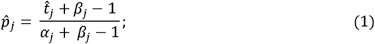

where 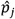 is the adjusted prevalence of disease (diabetes/hypertension) in the age group *j*,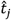 is the prevalence of disease estimated in the data for the age group *j*, and *α*_*j*_ and *β*_*j*_ are the sensitivity and specificity of the self-reported information in the *j* -*th* age group (Table 1). Rogan and Gladen (1978) showed that 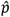 is unbiased when *α* and *β* are known, noticing that 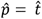 if and only if *α* = *β* = 1, i.e., when the test is perfect. Table 2 presents the prevalence of diabetes and hypertension before and after applying the adjustment formula.

**Table 2.** Prevalence of diabetes and hypertension for age groups adjusted and not adjusted Diabetes Hypertension.

| Age group | Diabetes |  | Hypertension |  |
| --- | --- | --- | --- | --- |
|  | Not adjusted (%) | Adjusted (%) | Not adjusted (%) | Adjusted (%) |
| 20-39 years | 2.44 | 3.97 | 3.81 | 7.83 |
| 40-60 years | 14.96 | 22.49 | 19.10 | 30.88 |
| 60 years or older | 32.79 | 42.88 | 44.68 | 69.37 |

We used a sample balancing method “*raking”* to replicate the adjusted prevalence of diabetes and hypertension. Raking is a statistical method that adjusts a set of data so that its marginal totals match control totals [20]. To apply raking, first, we considered that all subjects in the sample had the same weight (=1). Being diabetes and hypertension the auxiliary variables, we estimated the adjusted totals by age group, as follows:

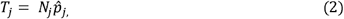

where *N*_*j*_ is the number of people in the age group j. Raking involves iteratively stratifying the sample using the set of auxiliary variables, and adjusting the weights to match the totals from the sample *N*_*j*_) with the totals provided (*T*_*j*_). We could use prior methods of misclassification analysis [21], but raking allows a faster way to estimate risk ratios and attributable fractions. Weight calibration was performed using a survey package on R Statistical Software [22–24]. The estimated weights were calculated after eliminating missing data for diabetes, obesity and hypertension, and eliminating subjects under 20 years old, but before any other exclusions.

### Covariates

Covariates included age (years), sex (male/female), state of residence (Mexico City as reference), cases recorded by type of epidemiological surveillance system (sentinel/no sentinel), health system and delay in seeking medical attention. We included sentinel/no sentinel as a potential confounder, given the differences in COVID-19 testing procedures. The health system was divided into five categories: 1) Mexican Institute of Social Security which provides care for formal workers (IMSS, by its acronym in Spanish), 2) Institute of Security and Social Services for State Workers that cares for State-affiliated employees (ISSSTE, by its acronym in Spanish), 3) private healthcare, 4) Health Secretary (SSA, by its acronym in Spanish) and IMSS *bienestar* that provide care for the uninsured population, and 5) Other health systems such as the Red Cross, National System for the Integral Development of the Family (DIF, by its acronym in Spanish), Mexican Petroleum’s Health Services (PEMEX, by its acronym in Spanish), National Defense Secretary (SEDENA, by its acronym in Spanish), Marine Secretary (SEMAR, by its acronym in Spanish), and University-based systems; under this label we also included individuals with missing data (n=10). Finally, we included delay in seeking medical attention (days) to capture the delay in receiving medical attention since the beginning of symptoms.

### Statistical analysis

#### Descriptive analysis

Descriptive statistics were used to estimate the prevalence of sociodemographic characteristics and multimorbidity; mean and 95% confidence intervals were used for delay in seeking medical attention. Analyses were stratified by age group: 20 to 39, 40 to 59, and 60 years or older.

#### Risk estimation

To estimate the risk of hospitalization and death, Poisson regression models with robust standard errors were implemented. We adjusted each model for the main confounders: age, sex, state of residence (Mexico City as reference), type of surveillance system (sentinel/hospital), health system (IMSS/ ISSSTE/ private, SSA and IMSS welfare, other health system) and delay in seeking medical attention. We considered the survey weights in Stata 14.0 to adjust for misclassification bias due to undiagnosed cases of diabetes and hypertension (College Station, TX) [25].

#### Estimation of the population attributable fraction (PAF)

We estimated the proportion of hospitalizations and deaths attributable to chronic diseases, using the Population Attributable Fraction (PAF) [26,27]:

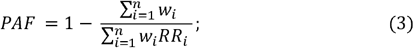

where *w*_*l*_ and *RR*_*l*_ denote the sampling weight and relative risk of the *i*-th individual in the sample.

### Sensitivity analyses

Sensitivity analyses were conducted to assess the impact of misclassification bias by comparing our main scenario to the scenario with no misclassification adjustment for diabetes and hypertension. Also, we assessed the impact of varying the number of diseases included in the multimorbidity variable by including: 1) three diseases: obesity, hypertension and diabetes; 2) nine diseases: obesity, COPD, hypertension, diabetes, cardiovascular disease, chronic kidney disease, asthma, immunodeficiency, and other comorbidities.

## Results

Table 3 presents the sociodemographic characteristics of COVID-19 positive adult patients by age group. The proportion of men increased with age. Obesity, hypertension and diabetes were the most frequent chronic diseases in all age groups. In adults aged 20-39 years, obesity (15.5%) and hypertension (8.9%) were the most prevalent diseases. Adults with 40-59 presented a higher prevalence of hypertension (33.8%) then diabetes (25.9%), and then obesity (22.7%). Individuals with no chronic diseases decreased from 76.2% among 20-39 yrs, to 46.5% among 40-59 yrs and to 18% among 60 yrs or more. In contrast, individuals with three or more diseases increased with age: from 1.3% among 20-39 yrs to 17.8% among 60 yrs and older.

**Table 3.**
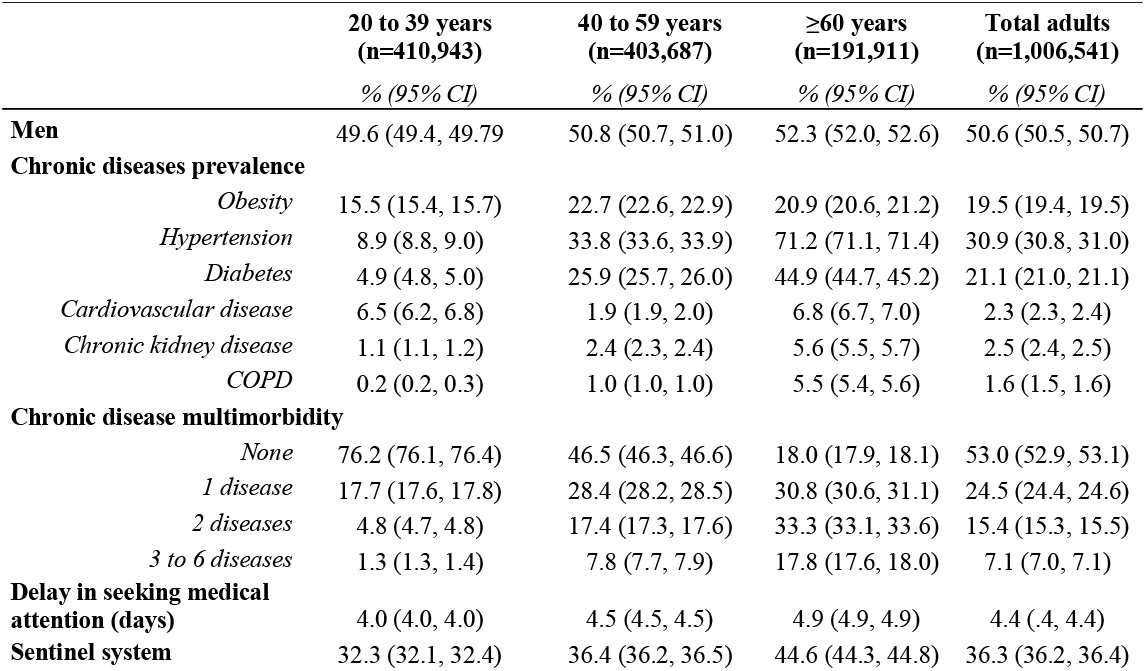
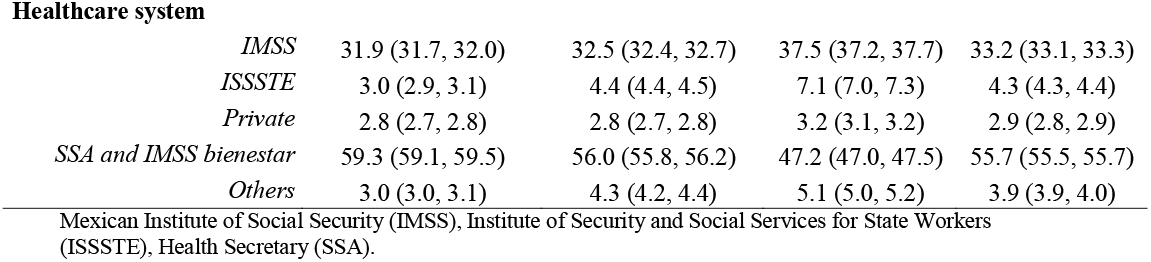
Demographic characteristics of COVID-positive cases in Mexico by age group.

|  | **20 to 39 years**<br>**(n=410,943)** | **40 to 59 years**<br>**(n=403,687)** | **≥60 years**<br>**(n=191,911)** | **Total adults**<br>**(n=1,006,541)** |
| --- | --- | --- | --- | --- |
|  | *% (95% CI)* | *% (95% CI)* | *% (95% CI)* | *% (95% CI)* |
| **Men** | 49.6 (49.4, 49.79) | 50.8 (50.7, 51.0) | 52.3 (52.0, 52.6) | 50.6 (50.5, 50.7) |
| **Chronic diseases prevalence** |  |  |  |  |
| *Obesity* | 15.5 (15.4, 15.7) | 22.7 (22.6, 22.9) | 20.9 (20.6, 21.2) | 19.5 (19.4, 19.5) |
| *Hypertension* | 8.9 (8.8, 9.0) | 33.8 (33.6, 33.9) | 71.2 (71.1, 71.4) | 30.9 (30.8, 31.0) |
| *Diabetes* | 4.9 (4.8, 5.0) | 25.9 (25.7, 26.0) | 44.9 (44.7, 45.2) | 21.1 (21.0, 21.1) |
| *Cardiovascular disease* | 6.5 (6.2, 6.8) | 1.9 (1.9, 2.0) | 6.8 (6.7, 7.0) | 2.3 (2.3, 2.4) |
| *Chronic kidney disease* | 1.1 (1.1, 1.2) | 2.4 (2.3, 2.4) | 5.6 (5.5, 5.7) | 2.5 (2.4, 2.5) |
| *COPD* | 0.2 (0.2, 0.3) | 1.0 (1.0, 1.0) | 5.5 (5.4, 5.6) | 1.6 (1.5, 1.6) |
| **Chronic disease multimorbidity** |  |  |  |  |
| *None* | 76.2 (76.1, 76.4) | 46.5 (46.3, 46.6) | 18.0 (17.9, 18.1) | 53.0 (52.9, 53.1) |
| *1 disease* | 17.7 (17.6, 17.8) | 28.4 (28.2, 28.5) | 30.8 (30.6, 31.1) | 24.5 (24.4, 24.6) |
| *2 diseases* | 4.8 (4.7, 4.8) | 17.4 (17.3, 17.6) | 33.3 (33.1, 33.6) | 15.4 (15.3, 15.5) |
| *3 to 6 diseases* | 1.3 (1.3, 1.4) | 7.8 (7.7, 7.9) | 17.8 (17.6, 18.0) | 7.1 (7.0, 7.1) |
| **Delay in seeking medical attention (days)** | 4.0 (4.0, 4.0) | 4.5 (4.5, 4.5) | 4.9 (4.9, 4.9) | 4.4 (.4, 4.4) |
| **Sentinel system** | 32.3 (32.1, 32.4) | 36.4 (36.2, 36.5) | 44.6 (44.3, 44.8) | 36.3 (36.2, 36.4) |
| Healthcare system |  |  |  |  |
| *IMSS* | 31.9 (31.7, 32.0) | 32.5 (32.4, 32.7) | 37.5 (37.2, 37.7) | 33.2 (33.1, 33.3) |
| *ISSSTE* | 3.0 (2.9, 3.1) | 4.4 (4.4, 4.5) | 7.1 (7.0, 7.3) | 4.3 (4.3, 4.4) |
| *Private* | 2.8 (2.7, 2.8) | 2.8 (2.7, 2.8) | 3.2 (3.1, 3.2) | 2.9 (2.8, 2.9) |
| *SSA and IMSS bienestar* | 59.3 (59.1, 59.5) | 56.0 (55.8, 56.2) | 47.2 (47.0, 47.5) | 55.7 (55.5, 55.7) |
| *Others* | 3.0 (3.0, 3.1) | 4.3 (4.2, 4.4) | 5.1 (5.0, 5.2) | 3.9 (3.9, 4.0) |

Mexican Institute of Social Security (IMSS), Institute of Security and Social Services for State Workers (ISSSTE), Health Secretary (SSA).

Table 4 presents the risk of hospitalization and death among individuals by multimorbidity status and age group. Adults under 40 years with three or more diseases presented 5.1 times higher risk of hospitalization (95% CI: 4.8, 5.4) and 15.2 times higher risk of death (95% CI: 13.5, 17.2), compared to disease-free individuals of the same age group. Adults aged 40-59 years with three or more diseases compared to individuals from the same age group with no diseases, presented 2.3 times higher risk of hospitalization (95% CI: 2.3, 2.4) and 3.7 times higher risk of death (95% CI: 3.6, 3.8). Adults aged 60-years or older with three or more diseases presented 36% higher hospitalization risk (95% CI: 1.3, 1.4) and 1.5 times higher risk of death (95% CI: 1.47, 1.54), compared to disease-free individuals from the same age group.

**Table 4.**
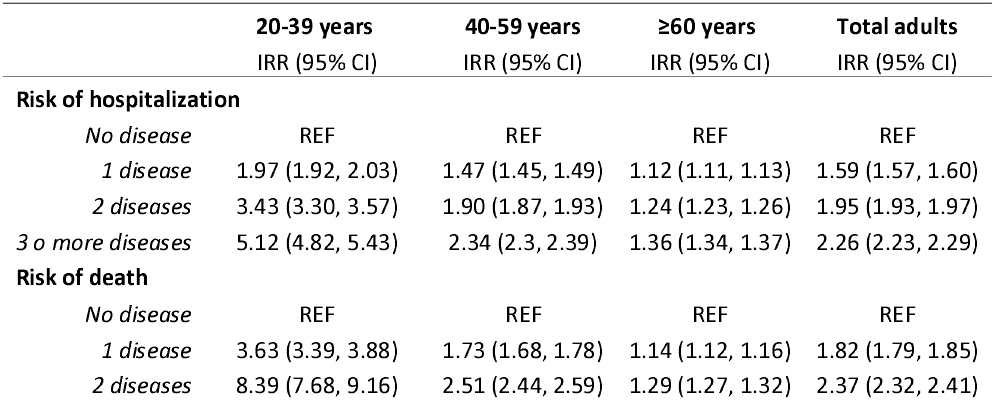
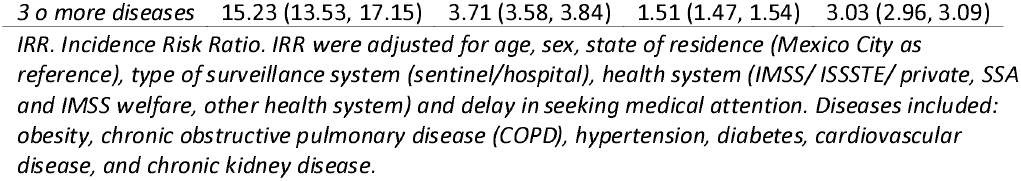
Risk of hospitalization and death among individuals with chronic diseases by age group.

Fig 1 shows the fraction of hospitalizations and deaths from COVID-19 attributable to chronic diseases. Hospitalizations and deaths attributable to chronic diseases were higher in younger than older adults. In adults under 40 years, 25.4% of hospitalizations (95% CI: 24.8%, 26.1%), and 50.1% of deaths (95% CI: 48.6%, 51.5%) could have been avoided by preventing the six diseases associated with poor nutrition and smoking. In adults aged 40-59 years, chronic diseases contributed to 28.3% of hospitalizations (95% CI: 27.8%, 28.7%) and 40.5% of deaths (95% CI: 39.7%, 41.3%), while for adults 60 years and older, 15.3% of hospitalizations (95% CI: 14.9%, 15.7%) and 18.7% of deaths (95% CI: 18.0%, 19.5%) could have been avoided.

**Fig 1.**
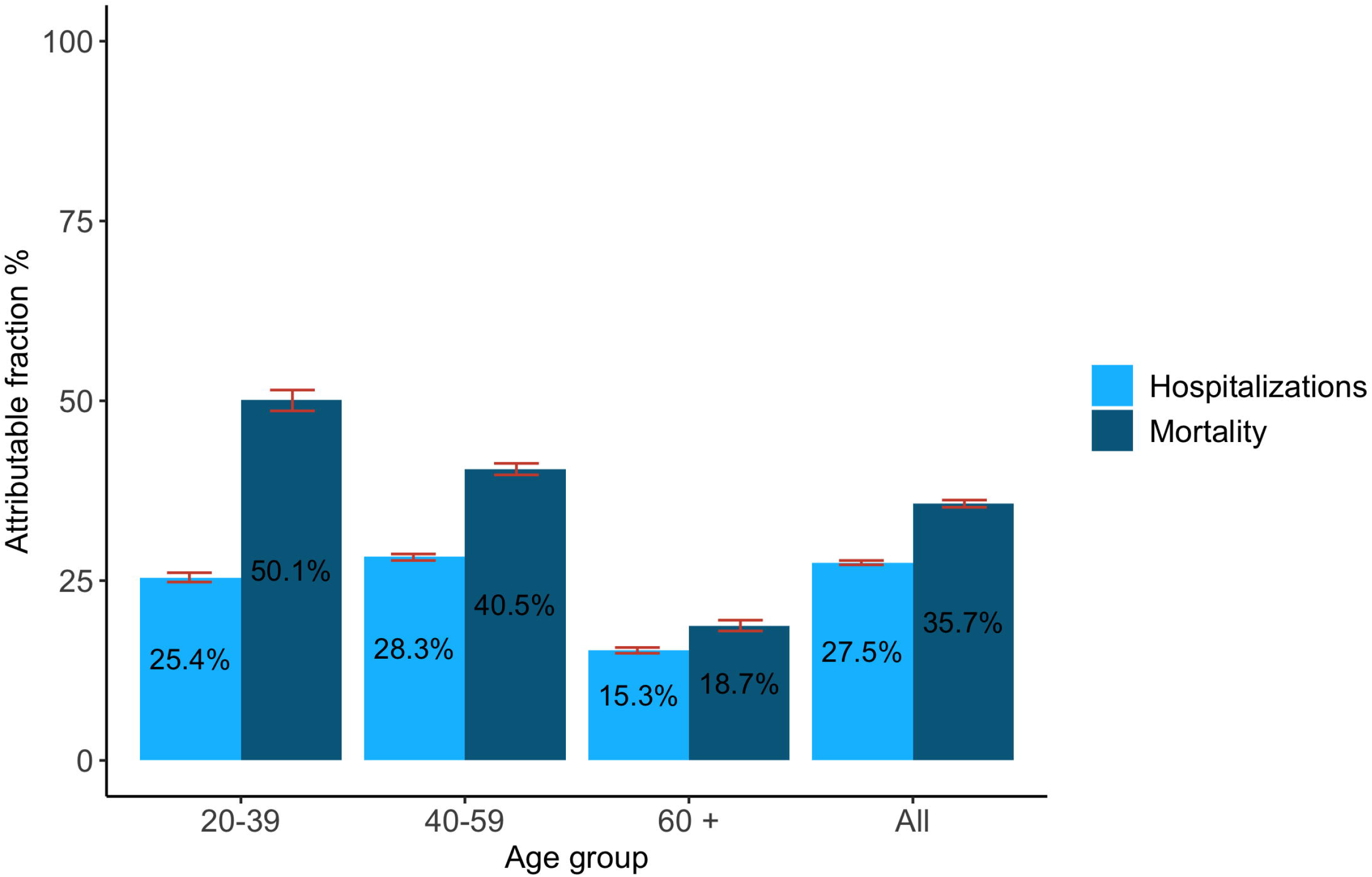
Fraction of hospitalizations and deaths from COVID-19 attributable to chronic diseases associated with poor nutrition and smoking.

Table 5 describes the sensitivity analyses conducted to understand the uncertainty around our estimates. First, estimates calculated without adjusting for misclassification bias in diabetes and hypertension were smaller than our main scenario. Across all age groups and considering the six chronic diseases, our main scenario estimates that 27.5% of all hospitalizations and 35.7% of all deaths were attributable to chronic diseases; yet, these estimates would have been 18.5% and 23.2% if unadjusted. The impact of misclassification adjustment was stronger for younger age groups. Second, by varying our definition of multimorbidity we observed that the burden for hospitalization and mortality is mainly explained by obesity, hypertension and diabetes: 27.5% of hospitalizations and 35.7% of deaths could be averted by preventing all six diseases associated with malnutrition and tobacco, yet 26.0% and 33.6% would have been prevented if we could have only prevented obesity, hypertension and diabetes. When considering all nine chronic diseases, the PAF slightly increased for hospitalization (all ages: from 27.5% to 28.7%) and mortality (all ages: from 35.7% to 37.3%). Relative risk estimates for each analyzed scenario are presented in the S1 Table.

**Table 5.**
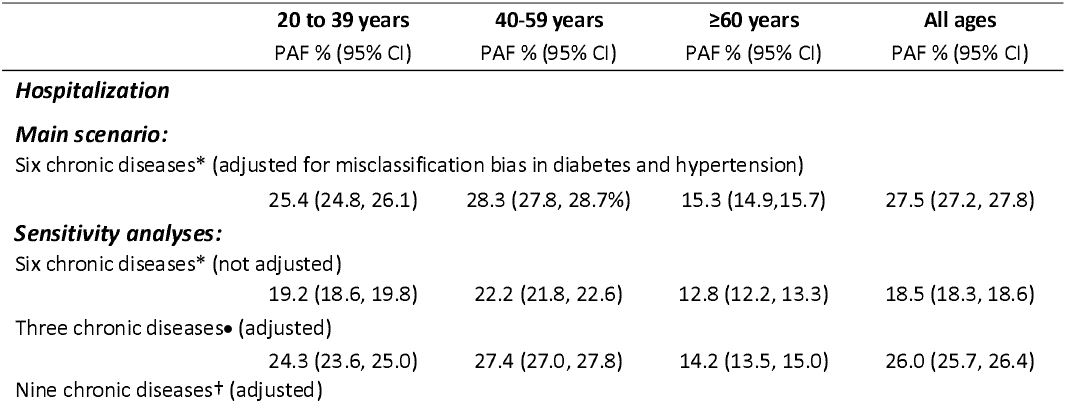
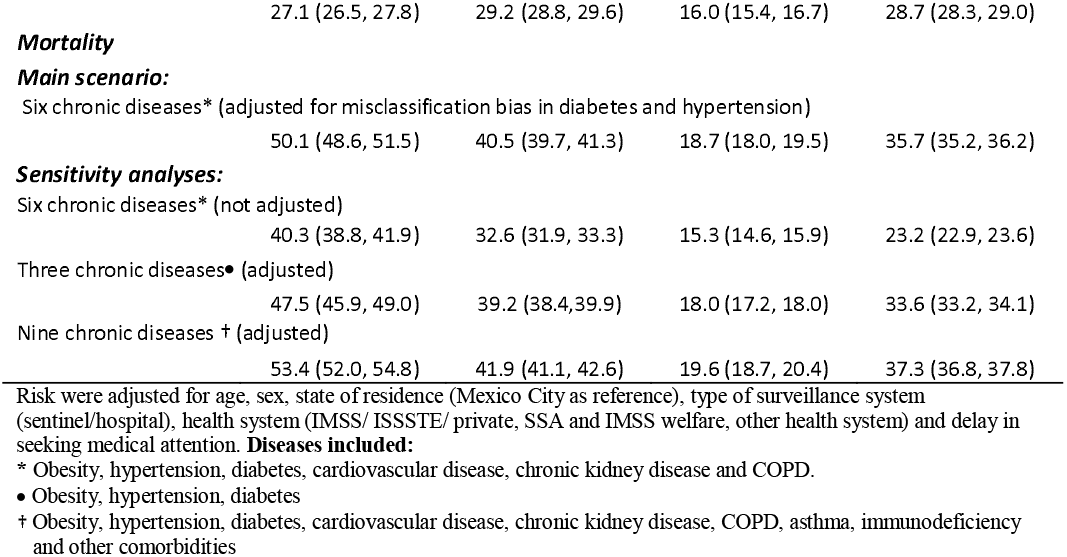
Sensitivity analyses: Fraction of hospitalization and deaths attributable to chronic diseases under different scenarios.

## Discussion

We aimed to estimate the proportion of hospitalizations and deaths from COVID-19 attributable to chronic diseases (PAF). The six chronic diseases analyzed contributed to 28% of hospitalizations and 36% of deaths from COVID-19 among Mexican adults. The burden of chronic diseases on COVID-19 was higher in younger than older adults: 25% of hospitalizations and 50% of deaths could have been averted by preventing chronic diseases among 20-39 yrs versus 15% of hospitalizations and 19% of deaths among adults 60 yrs and older.

In Mexico, one study estimated the PAF on COVID-19 mortality using data of the social security institution (IMSS) [5]. They estimated that 2% of deaths from COVID-19 were attributed to diabetes, 7% to hypertension, and 8% to obesity among inpatients. This proportion changed to 1.1% 14.3% and 16.8%, respectively, among outpatients. This study did not attempt to estimate the combined effect of chronic diseases, so no direct comparison is possible. In a supplementary analysis, we stratified our models by patients treated as inpatients or outpatients (S2 Table) and found a higher proportion of deaths attributable to chronic diseases among outpatient (35.3%; 95%CI: 33.9, 36.7) than inpatients (19.8%; 95% CI: 19.0, 20.6). The difference could be explained by an overrepresentation of individuals with chronic diseases among inpatients compared to outpatients - people with chronic diseases are more likely to be hospitalized, even with mild symptoms - leading to an underestimation of risk that is not observed among outpatients [28]. Our analysis included a multimorbidity approach that enabled us to estimate the overall burden of chronic diseases, a longer time frame, and an adjustment for misclassification bias for diabetes and hypertension. Despite differences in methodology, both studies suggest that chronic diseases have a substantial impact on COVID-19 deaths.

The proportion of severe COVID-19 attributable to multimorbidity has been previously studied in other countries. In the United Kingdom, 51% of severe COVID-19 cases were attributable to unhealthy behaviors (13% to smoking, 9% to physical inactivity, and 30% to overweight and obesity) [8]. In the US, 36% of severe COVID-19 cases were attributable to unhealthy behaviors (14% to smoking, 12% to physical activity, and 16% to diet) [9]. In another US study, 63% of hospitalizations were attributable to the jointly effect of diabetes, obesity, hypertension and heart failure. In our study, 27.5% of COVID-19 hospitalizations were attributable to six chronic diseases associated with smoking and poor nutrition. The PAF in Mexico was smaller than in the UK and US. Estimates in the UK and US include different risk factors and different methodologies to combine risks, which could explain some differences. We employed a multimorbidity approach to combine diseases and avoid double counting.

One study in Mexico used a multimorbidity perspective to estimate the risk of mortality from COVID [29]. Authors found that adults aged 20-39 years with three chronic diseases presented 16.1 (95% CI: 13.8, 18.7) times higher risk of death, adults aged 40-59 years presented 4.2 (95% CI: 4.0, 4.4) and adults 60-80 years 1.6 (95% CI:1.6 1.7), compared to disease-free individuals of the same age group. Their results were consistent with ours: 15.2 (95% CI: 13.5, 17.15), 3.7 (95% CI: 3.6, 3.8) and 1.5 (95% CI: 1.5, 1.5) for adults aged 20-39, 40-59 and ≥60, respectively. These results are consistent with our estimations. Another study in Mexico estimated that obesity mediates half of the diabetes effect in the risk of dying from COVID-19 [30]. This study shows that diseases are not independent and highlights the need of using a multimorbidity strategy to evaluate the burden of chronic diseases on COVID. In our study, having a single chronic disease increased 82% the risk of dying from COVID-19, but the risk increased 2.4 times if the patient had two diseases and 3 times with three or more diseases.

Studies in other countries showed the need to use a multimorbidity approach on COVID. Studies found that patients with multimorbidity have a higher risk of COVID-19 complications than those having a single disease. In Italy, a retrospective study found higher odds of severity in patients with cardiometabolic multimorbidity, compared to patients with no cardiometabolic condition (OR: 3.19; 95%CI 1.61–6.34) [31]. Also, patients with diabetes showed higher odds of severity from COVID-19, compared with patients without diabetes, but the magnitude of the association was mainly driven by multimorbidity rather than by diabetes alone. Another study in Italy using a comorbidity index, including diabetes, cardiovascular disease, chronic kidney disease among others, showed that non-survivors to COVID-19 had a higher comorbidity score than survivors [32]. The higher risk of complications in patients with multimorbidity in comparison with a single cardiometabolic disease implies that studies need to consider the cardiometabolic risk as a whole, and not each disease individually.

The data used in our analysis was produced in the context of the epidemiological surveillance system of respiratory diseases in Mexico and is subject to many sources of bias. First, our database lacks follow-up: the data is collected when the patient is admitted to the medical facility, and only death has a longitudinal follow-up. Second, data on chronic diseases is self-reported and a large proportion of the population in Mexico with chronic diseases is undiagnosed. We used a bias quantification approach to adjust our estimates, producing an increase in the PAF from 18.5% to 27.5% for hospitalization and from 23.3% to 35.7% in deaths (Table 5). Unfortunately, we did not have information to adjust the misclassification bias of the other four diseases considered in our main analysis; in particular, we think the adjustment of obesity could have further increase the PAF, considering its large impact on COVID-19 severity and mortality and its high prevalence in the Mexican population, thus, we are still likely underestimating the PAF. We did not have information about chronic disease control, which could further increase mortality [33,34]. Future studies should focus on chronic disease control, ideally using well-defined denominators taking advantage of data collected in prior cohort studies. Our study could also be subject to selection bias. Our sample is composed mainly by subjects with severe COVID-19, since all severe cases of respiratory disease are subject to the COVID-19 test, while only a subsample of mild cases is tested. Then, the global association captures both the causal factors of COVID-19 and the imbalance in access to RT-PCR; unfortunately, we did not have access to an estimate of the total number of mild respiratory disease, which could have allowed us to produce a better estimate. Also, we do not know the mortality coverage of the surveillance system. The mortality registry in Mexico has important delays, and a significant proportion of deaths have not been reported in the official COVID-19 registry, as evidenced by excess mortality estimates [35]. Hospital deaths are often registered quicker in the COVID-19 dataset, while home deaths or people who died without being tested are not be updated at the same speed, or in some cases not reported. It is difficult to predict the direction of the bias, as it will depend on whether the differential registry is informed by chronic diseases.

Latin America is one of the regions more heavily affected by COVID-19 mortality in young adults. Developing countries show a clearly different mortality pattern than developed countries, where young adult mortality is rare [36]. In our study, we found that up to 50% of the COVID-19 mortality burden is attributable to chronic diseases in younger adults, compared to 18.7% in adults 60 years of age and older. This allows us to understand the relative importance of chronic disease in young adult’s mortality, yet, it also shows that a large proportion of the mortality is attributable to other causes. A recent study showed that the differential burden of mortality in young adults between developed and developing countries is attributable to a higher infection rates among young adults in developing countries, but also to a lower recovery rate once infected, driven by chronic diseases and poorer healthcare access [36]. We did not explore healthcare access in our analysis, and that could certainly contribute to higher mortality. Future studies should explore other causes linked to increased mortality, such as quality of care.

## Conclusion

We found that multimorbidity associated with poor nutrition produced a sizable proportion of the hospitalizations and deaths from COVID-19, particularly among young adults in Mexico. Mexico has been dealing with the double challenge of high demand for hospital services due to COVID-19 and treating chronic diseases. Individual efforts to control and reduce chronic diseases are direly needed in the short term. In the longer term, implementing structural interventions such as taxes on tobacco, sugary beverages, and high-energy foods of low nutritional value, warning labels, prohibition of advertisements to non-nutritional food and beverages and smoke-free spaces will be critical to reduce the burden of chronic diseases and COVID-19.

## Data Availability

The data that support the findings of this study are openly available at http://www.gob.mx/salud/documentos/datos-abiertos-152127

