## Supplementary material for "Fraction of COVID-19 hospitalizations and deaths attributable to chronic diseases": S1 Table

**S1 Table.** **Sensitivity analyses: Risk of hospitalization and risk of death among individuals with chronic diseases by age groups and total adults under different scenarios**

|  | **20 to 39 years** | **40-59 years** | **≥60 years** | **Total adults** |
| --- | --- | --- | --- | --- |
|  | IRR (95% CI) | IRR (95% CI) | IRR (95% CI) | IRR (95% CI) |
| Hospitalization |  |  |  |  |
| corrected for misclassification bias | |  |  |  |
| *No disease* | REF | REF | REF | REF |
| *1 disease* | 1.97 (1.92, 2.03) | 1.47 (1.45, 1.49) | 1.12 (1.11, 1.13) | 1.59 (1.57, 1.6) |
| *2 diseases* | 3.43 (3.3, 3.57) | 1.9 (1.87, 1.93) | 1.24 (1.23, 1.26) | 1.95 (1.93, 1.97) |
| *3 o more diseases* | 5.12 (4.82, 5.43) | 2.34 (2.3, 2.39) | 1.36 (1.34, 1.37) | 2.26 (2.23, 2.29) |
| Not corrected for misclassification bias | | |  |  |
| *No disease* | REF | REF | REF | REF |
| *1 disease* | 1.95 (1.9, 2) | 1.95 (1.9, 2) | 1.15 (1.13, 1.16) | 1.48 (1.47, 1.5) |
| *2 diseases* | 3.38 (3.26, 3.51) | 3.38 (3.26, 3.51) | 1.25 (1.24, 1.26) | 1.74 (1.73, 1.76) |
| *3 o more diseases* | 5.01 (4.72, 5.32) | 5.01 (4.72, 5.32) | 1.36 (1.34, 1.37) | 1.97 (1.95, 1.99) |
| Obesity, diabetes and hypertension (corrected) | | |  |  |
| *No disease* | REF | REF | REF | REF |
| *1 disease* | 2.1 (2.04, 2.15) | 1.48 (1.46, 1.5) | 1.12 (1.1, 1.13) | 1.58 (1.56, 1.59) |
| *2 diseases* | 3.12 (2.99, 3.26) | 1.94 (1.91, 1.97) | 1.25 (1.23, 1.26) | 1.92 (1.9, 1.94) |
| *3 o more diseases* | 4.45 (4.11, 4.82) | 2.19 (2.14, 2.24) | 1.34 (1.32, 1.36) | 2.21 (2.18, 2.24) |
| 9 chronic diseases (corrected) | |  |  |  |
| *No disease* | REF | REF | REF | REF |
| *1 disease* | 1.91 (1.86, 1.96) | 1.46 (1.44, 1.48) | 1.12 (1.11, 1.13) | 1.58 (1.56, 1.59) |
| *2 diseases* | 3.23 (3.11, 3.35) | 1.89 (1.86, 1.92) | 1.25 (1.23, 1.26) | 1.96 (1.94, 1.98) |
| *3 o more diseases* | 5.1 (4.85, 5.38) | 2.34 (2.3, 2.38) | 1.36 (1.34, 1.37) | 2.27 (2.24, 2.3) |
| Death |  |  |  |  |
| corrected for misclassification bias | |  |  |  |
| *No disease* | REF | REF | REF | REF |
| *1 disease* | 3.63 (3.39, 3.88) | 1.73 (1.68, 1.78) | 1.14 (1.12, 1.16) | 1.82 (1.79, 1.85) |
| *2 diseases* | 8.39 (7.68, 9.16) | 2.51 (2.44, 2.59) | 1.29 (1.27, 1.32) | 2.37 (2.32, 2.41) |
| *3 o more diseases* | 15.23 (13.53, 17.15) | 3.71 (3.58, 3.84) | 1.51 (1.47, 1.54) | 3.03 (2.96, 3.09) |
| Not corrected for misclassification bias | | |  |  |
| *No disease* | REF | REF | REF | REF |
| *1 disease* | 3.57 (3.34, 3.81) | 1.76 (1.71, 1.8) | 1.17 (1.15, 1.19) | 1.61 (1.59, 1.64) |
| *2 diseases* | 8.05 (7.39, 8.76) | 2.47 (2.4, 2.55) | 1.3 (1.28, 1.33) | 1.99 (1.95, 2.02) |
| *3 o more diseases* | 14.69 (13.08, 16.5) | 3.62 (3.5, 3.75) | 1.5 (1.47, 1.54) | 2.48 (2.43, 2.53) |
| Obesity, diabetes and hypertension (corrected) | | |  |  |
| *No disease* | REF | REF | REF | REF |
| *1 disease* | 3.97 (3.72, 4.24) | 1.75 (1.7, 1.8) | 1.15 (1.13, 1.17) | 1.79 (1.76, 1.82) |
| *2 diseases* | 7.61 (6.94, 8.36) | 2.63 (2.56, 2.71) | 1.32 (1.3, 1.34) | 2.35 (2.31, 2.4) |
| *3 o more diseases* | 10.22 (8.65, 12.06) | 3.31 (3.17, 3.45) | 1.51 (1.47, 1.55) | 2.93 (2.86, 3) |
| 9 chronic diseases (corrected) | |  |  |  |
| *No disease* | REF | REF | REF | REF |
| *1 disease* | 3.55 (3.32, 3.81) | 1.73 (1.68, 1.78) | 1.15 (1.13, 1.17) | 1.83 (1.8, 1.86) |
| *2 diseases* | 8.15 (7.47, 8.9) | 2.5 (2.42, 2.58) | 1.3 (1.27, 1.32) | 2.39 (2.34, 2.43) |
| *3 o more diseases* | 15.45 (13.89, 17.19) | 3.68 (3.56, 3.81) | 1.49 (1.46, 1.52) | 3.03 (2.96, 3.09) |
