## Supplementary material for "Fraction of COVID-19 hospitalizations and deaths attributable to chronic diseases": S2 Table

Table 2 presents the risks of death and the fraction of deaths attributable to chronic diseases among outpatients and inpatients with chronic diseases. The risk of dying from COVID-19 in people with chronic diseases was higher among outpatients adults than inpatients adults and higher among those with three or more diseases. Outpatients with 3 or more chronic diseases presented 4.29 times (95% CI: 4.0, 4.60) greater risk of death compared to disease free individuals of the same group. Inpatients with 3 or more diseases compared to individuals from the same group with no diseases, presented 1.5 times (95% CI: 1.47, 1.53) higher risk of death. Deaths attributable to chronic diseases were higher in outpatients than inpatients. In inpatients, 35.3% (95% CI: 33.9%, 36.7%) of deaths could have been avoided by preventing the six diseases associated with poor nutrition and smoking. In outpatients, chronic diseases contributed to 19.8% (95% CI: 19.0%, 20.6%) of deaths.

**S2 Table.** **Risks of death and attributable fractions estimated among outpatients and inpatients with chronic diseases.**

|  | **N (%)** | **Mortality** | **RR (95% CI)** | **PAF % (95% CI)** |
| --- | --- | --- | --- | --- |
| ***Outpatiens*** |  |  |  |  |
| **Total** | **786343.34 (100)** | **11209** | **-** | **35.3 (33.9, 36.7)** |
| No disease | 482814 (61.4) | 2212 | 1.00 |  |
| 1 disease | 182929 (23.2) | 3184 | 1.90 (1.80, 2.00) |  |
| 2 diseases | 88494 (11.3) | 3476 | 2.79 (2.63, 2.97) |  |
| 3 o more diseases | 32107 (4.1) | 2336 | 4.29 (4.00, 4.60) |  |
| ***Inpatients*** |  |  |  |  |
| **Total** | **237351 (100)** | **91734** | **-** | **19.8 (19.0, 20.6)** |
| No disease | 60111 (25.3) | 15704 | 1.00 |  |
| 1 disease | 68125 (28.7) | 25940 | 1.22 (1.20, 1.24) |  |
| 2 diseases | 68797 (29.0) | 30295 | 1.34 (1.31, 1.36) |  |
| 3 o more diseases | 40319 (17.0) | 19796 | 1.50 (1.47, 1.53) |  |

**RRs were estimated using the sampling weights obtained via calibration (Raking).*

**** *These PAFs cannot be added because each of them was estimated in different populations: (1) outpatients or (2) inpatiens.*
